# Prevalence of intraventricular haemorrhage and determinants of its early outcomes among preterm neonates at the newborn unit of a teaching hospital in Western Kenya

**DOI:** 10.1101/2022.03.09.22272142

**Authors:** Gloria Namubuya Sisenda, Festus Muigai Njuguna, Winstone Mokaya Nyandiko

**Author notes:** **Corresponding Author:** Dr. Gloria Namubuya Sisenda, Department of Child health and Paediatrics, Moi University College of Health Sciences, P.O Box 4606-30100, Eldoret-Kenya.

## Abstract

Intraventricular haemorrhage (IVH) screened using cranial ultrasounds (cUS) is a major cause of morbidity and mortality among preterm neonates. Despite the adverse neonatal outcomes attributed to IVH, there are limited studies conducted in sub-Saharan Africa on IVH occurrence and determinants of early outcomes. This study assessed the proportion of neonates with IVH, its determinants and the early outcomes of preterm neonates with the condition. A prospective descriptive study conducted at the newborn unit of Moi Teaching and Referral Hospital in Western Kenya between March 2020 to March 2021. The neonates sampled systematically had their clinical characteristics and that of their mothers collected from medical records. A cUS screening was conducted on the third and fourteenth day of life while hydrocephalus was screened using serial weekly measurements of head circumference and mortality assessed within the first 28 days of life. Bivariate analysis was used to test for an association between patient characteristics, occurrence of IVH and early outcomes. Confounders were controlled using a multivariate logistic regression model. We enrolled 201 pre-term neonates of whom 105 (52.2%) were male and 68 (33.8%) had IVH. Among neonates with IVH, 46 (67.6%) had mild while 22 (32.4%) had severe IVH. Antenatal steroids significantly reduced the risk of IVH while extreme/very low gestational age and extremely low birthweight significantly increased the risk of IVH two and three-fold respectively. Neonates with thrombocytopenia and on mechanical ventilation were significantly more likely to be diagnosed with IVH. Early outcomes of IVH were hydrocephalus (4.0%) and mortality (19.1%). Intraventricular haemorrhage was seen in one-third of neonates enrolled with majority of them presenting with the lower IVH grades. The IVH risk significantly increased among neonates with very low gestational age and extremely low birthweight but was lowered by antenatal steroid use. Mortality rates were significantly higher among neonates with thrombocytopenia.

## Background

Preterm neonates with a very low birthweight are at risk of health problems at birth and mortality in the first four weeks of life [1]. For those who survive, there is an increased risk of long-term disabling neurological consequences or neurological problems that could interfere with achieving their developmental milestones and learning [2]. Several factors predispose these preterm neonates with very low birthweight to these complications. A major contributing factor to morbidity and mortality among one in five preterm newborns (below 32 weeks gestation) is intraventricular haemorrhage (IVH). This condition is multifactorial in nature and is often reported in cases of fibronectin deficiency in the germinal matrix basal lamina [3]. Intraventricular haemorrhage pathogenesis is associated with irregularities in cerebral blood flow, coagulation and platelet abnormalities that worsen the bleeding making this very critical infant in the integrity of the germinal matrix [4,5]. Some of the etiological risk factors for IVH include very low birthweight (<1500 grammes), very low to extremely low gestational age (< 32 weeks), absence of prenatal corticosteroid use for women at risk of premature mature delivery [2] and early clamping of the umbilical cord [6].

Prematurity is a leading cause of death in children below the age of five years with more than half of those born with a gestation age below 32 weeks dying in countries with low income such as those in the sub-Saharan Africa region [7]. With the high burden of prematurity, neonatal morbidity and infant mortality in countries with low- and middle-income economies, health conditions such as intraventricular haemorrhage further compound the problem in an already weak healthcare system. It is estimated that the neonatal mortality rate in sub-Saharan Africa is 27 per 1000 live births and 21 per 1000 live births in Kenya [8]. This is despite the United Nations sustainable development goal 3.2 which aims to reduce the neonatal mortality in all countries to 12 per 1000 live births [9]. Because of this and the high interest in child health evidenced by national strategies has led to an increased investment and development in neonatal care so as to improve survival rates [10]. The resulting high number of preterm, very low birth weight (VLBW) and extremely low birth weight (ELBW) newborns increases the risk of intracranial bleeding occurring, and intraventricular haemorrhage (IVH) in particular; with neonates born <32 weeks gestation age (GA) or <1500g being more susceptible [11,12]. While IVH contributes significantly to poor outcomes in mortality, morbidity, cognitive and motor neurological development, there is little data in its incidence, risk factors and complications in LICs as compared to high income countries [12]. This limited knowledge is in part because there have been few studies on IVH and its implications on the wellbeing of preterm and low birthweight newborns in sub-Saharan Africa. Furthermore, the cost of routine monitoring coupled with the systemic infrastructural inadequacy compound the problem further thus interfering with the ability to conduct a comprehensive surveillance [13]. Due to the heterogeneity of this demographic, perinatal exposures and difference in the quantity and quality of neonatal care between countries with developed and developing economies, the information from the former cannot be directly applied in understanding IVH implications on countries with developing economies such as Kenya. This study, therefore, hopes to contribute to the understanding of IVH and its implications in low income countries within the East Africa region and Western Kenya in particular. This was achieved by investigating the prevalence, factors associated and short-term outcomes of IVH among preterms in the newborn unit of a teaching hospital. We hope that the findings of this study will contribute to the body of knowledge that can lead to formulation of guidelines on screening for IVH in preterm neonates. This study therefore aimed to describe the prevalence, determinants and short-term outcomes of intraventricular haemorrhage (IVH) among preterm neonates in the new-born unit of a national teaching and referral hospital in Western Kenya. Specifically, it determined the proportion of preterm neonates with IVH, factors contributing to IVH among preterm neonates and the short-term (first 28 days) neonatal outcomes of those diagnosed with IVH at the newborn unit of Moi Teaching and Referral Hospital.

### Methodology

The study adopted a prospective descriptive study design at the newborn unit of Moi Teaching and Referral Hospital (MTRH) in Western Kenya between March 2020 to March 2021. The hospital is the second largest teaching and referral hospital in Kenya with a catchment area of the Western, North and South Rift regions. It has the largest newborn unit with highly skilled specialists in neonatology, paediatrics, neurology and neurosurgery; making it the most appropriate setting for a study on neonatal intraventricular haemorrhage. The neonates were sampled systematically (k=2) and both their clinical characteristics and that of their mothers collected from medical records. Each neonate then underwent cUS screening by a trained paediatric resident on the third and fourteenth day of life using a 6.5 MHz transducer and the cranial images reviewed by a radiologist. Serial weekly measurements of head circumference were then done to determine outcome of hydrocephalus and daily monitoring for occurrence of death within 28 days of life or on discharge. The data collected was analysed descriptively as proportions and a bivariate analysis was conducted to test for association between patient characteristics, occurrence of IVH and early outcomes using statistical package for social sciences (SPSS) version 24. Multivariate logistic regression was conducted to control for confounders if the bivariate analysis was statistically significant (p ≤ 0.05). Odds ratios were computed at 95% confidence interval. Written informed parental consents were obtained from all the mothers prior to enrolling their neonates. This study received ethical approval from the independent research and ethics committee (IREC) of Moi University School of Medicine and Moi Teaching and Referral Hospital (Approval number: 0001994).

## Results

This study enrolled 201 pre-term neonates of whom 105 (52.2%) were male. The mean birthweight of the neonates was 1251.9 (±185.4) grammes with 182 (90.6%) being between 1000-1500 grammes. The APGAR score was obtained at five minutes and 42 (20.9%) neonates had a low 5-minute APGAR score (below 7). Majority (60.2%) of the newborns had respiratory distress syndrome (RDS) and 116 (57.7%) were on mechanical ventilation. Nearly all (99%) of the newborns had a normal haemoglobin concentration (≥12 g/dl) and 178 (88.6%) had a normal platelet count (≥150000/µL) while 114 (56.7%) had a very low gestational age (<32 weeks) at the time of birth (Table 1).

**Table 1:** Neonatal characteristics.

| Variable | Category | Frequency (N) | Percentage (%) |
| --- | --- | --- | --- |
| Sex | Male | 105 | 52.2 |
|  | Female | 96 | 47.8 |
| Birth weight (grams) | <1000 | 19 | 9.4 |
|  | 1000-1500 | 182 | 90.6 |
|  | Mean (SD) | 1251.9 (185.4) |  |
| Apgar score (at 5 minutes) | ≤3 | 2 | 1.0 |
|  | 4 – 6 | 40 | 19.9 |
|  | 7 – 10 | 150 | 74.6 |
|  | Unknown | 9 | 4.5 |
| RDS | No | 80 | 39.8 |
|  | Yes | 121 | 60.2 |
| Platelets (#/μL) | ≥150000 (Normal) | 178 | 88.6 |
|  | <150000 (abnormal) | 23 | 11.4 |
| Hb (g/dl) | ≥12 (normal) | 199 | 99.0 |
|  | <12 (low) | 2 | 1.0 |
| GA (weeks) | < 28 | 3 | 1.5 |
|  | 28 – 31 | 111 | 55.2 |
|  | 32 – 34 | 87 | 43.3 |
| Mechanical ventilation | No | 85 | 42.3 |
|  | Yes | 116 | 57.7 |

The most common (39.8%) age group for the women whose newborns were enrolled in the study was 22-28 years and majority (54.2%; n=109) of all the new mothers had a parity of one. Only 43 (21.4%) used antenatal steroids during the antenatal period, 190 (94.5%) gave birth in a hospital setting of which 58 (28.9%) of all the births being caesarean sections (Table 2).

The prevalence of IVH among low-birthweight newborns at Moi Teaching and Referral Hospital in Western Kenya was 33.8% (n=68). Of these, 51.4% (n=35) were classified as Grade I using the Papile’s grading system for IVH; while there were equal proportions of newborns with Grades II, III and IV at 16.2% (n=11) each.

**Table 2:**
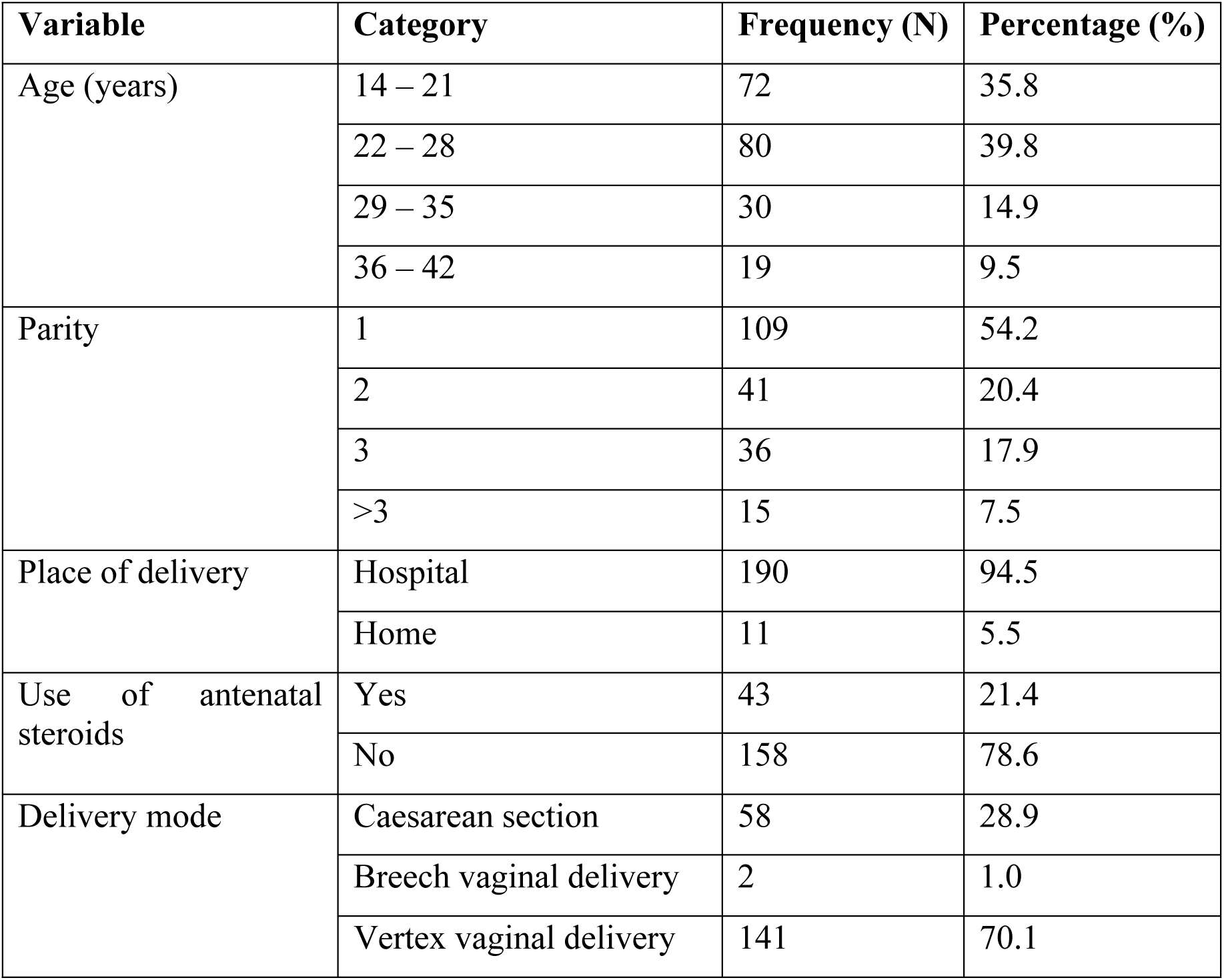
Maternal characteristics.

We report that the use of antenatal steroids significantly reduced the risk of IVH (RR=0.111; 95% CI: 0.028, 0.436; p<0.001) while extreme and very low gestational age below 32 weeks (RR=1.968; 95% CI: 1.255, 3.087; p=0.002) as well as an extremely low birthweight (RR=3.448; 95% CI:2.662, 4.467; p<0.001) significantly increased the risk of IVH two and three-fold respectively. We also noted that neonates on mechanical ventilation (RR=2.502; 95% CI: 1.658, 3.774; p<0.001) and had thrombocytopenia (RR=2.006; 95% CI: 1.350, 2.982; p=0.005) were significantly more likely to be diagnosed with IVH. However, there was no statistically significant association between maternal age, place of delivery and gender of baby with a diagnosis of intraventricular haemorrhage (Table 3).

**Table 3:**
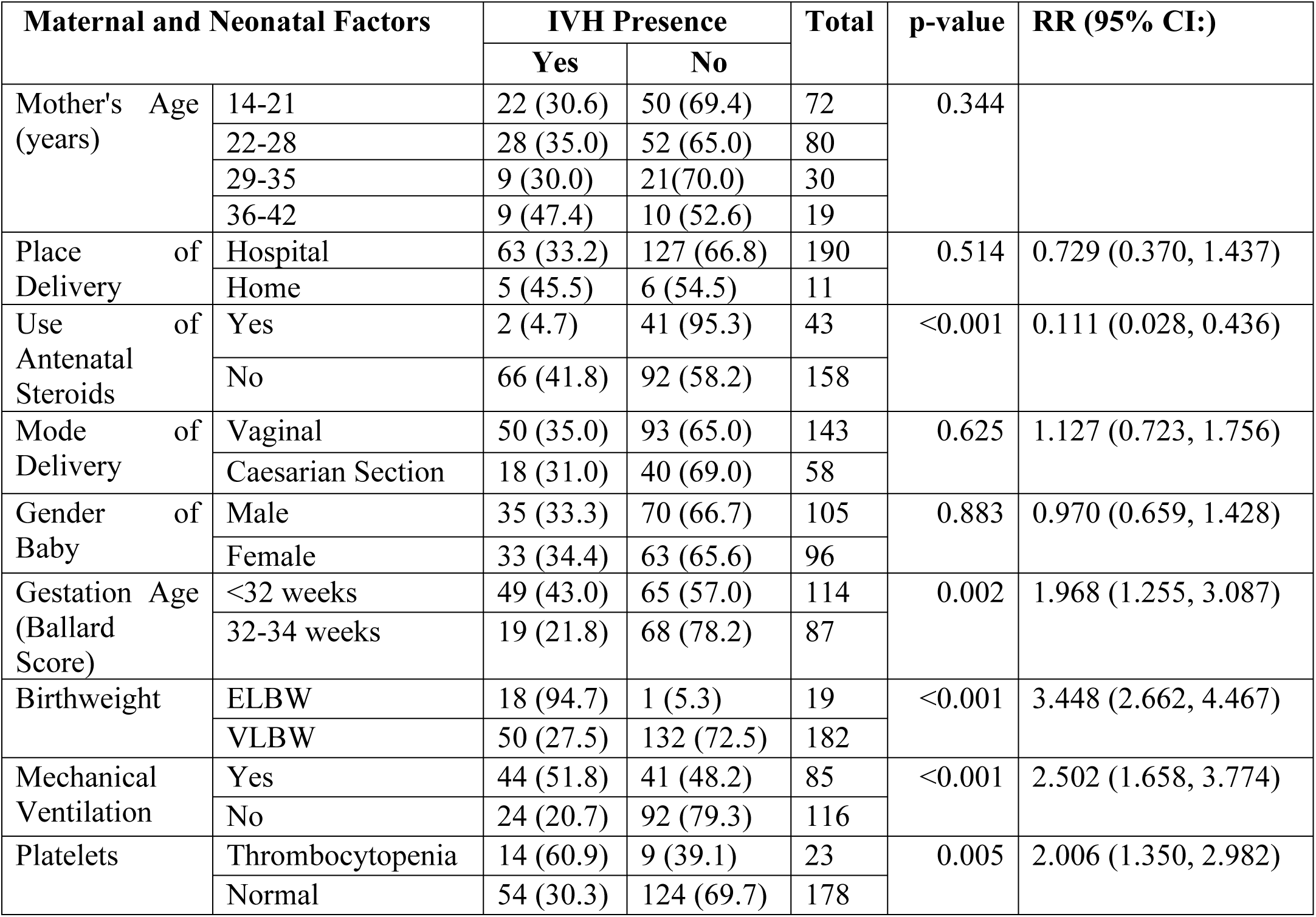
Maternal and Neonatal Factors Associated with intraventricular haemorrhage.

| Maternal and Neonatal Factors |  |  | IVH Presence |  | Total | p-value | RR (95% CI:) |
| --- | --- | --- | --- | --- | --- | --- | --- |
|  |  |  | Yes | No |  |  |  |
| Mother's Age (years) | 14-21 |  | 22 (30.6) | 50 (69.4) | 72 | 0.344 |  |
|  | 22-28 |  | 28 (35.0) | 52 (65.0) | 80 |  |  |
|  | 29-35 |  | 9 (30.0) | 21 (70.0) | 30 |  |  |
|  | 36-42 |  | 9 (47.4) | 10 (52.6) | 19 |  |  |
| Place of Delivery | Hospital |  | 63 (33.2) | 127 (66.8) | 190 | 0.514 | 0.729 (0.370, 1.437) |
|  | Home |  | 5 (45.5) | 6 (54.5) | 11 |  |  |
| Use of Antenatal Steroids | Yes |  | 2 (4.7) | 41 (95.3) | 43 | <0.001 | 0.111 (0.028, 0.436) |
|  | No |  | 66 (41.8) | 92 (58.2) | 158 |  |  |
| Mode of Delivery | Vaginal |  | 50 (35.0) | 93 (65.0) | 143 | 0.625 | 1.127 (0.723, 1.756) |
|  | Caesarian Section |  | 18 (31.0) | 40 (69.0) | 58 |  |  |
| Gender of Baby | Male |  | 35 (33.3) | 70 (66.7) | 105 | 0.883 | 0.970 (0.659, 1.428) |
|  | Female |  | 33 (34.4) | 63 (65.6) | 96 |  |  |
| Gestation Age (Ballard Score) | <32 weeks |  | 49 (43.0) | 65 (57.0) | 114 | 0.002 | 1.968 (1.255, 3.087) |
|  | 32-34 weeks |  | 19 (21.8) | 68 (78.2) | 87 |  |  |
| Birthweight | ELBW |  | 18 (94.7) | 1 (5.3) | 19 | <0.001 | 3.448 (2.662, 4.467) |
|  | VLBW |  | 50 (27.5) | 132 (72.5) | 182 |  |  |
| Mechanical Ventilation | Yes |  | 44 (51.8) | 41 (48.2) | 85 | <0.001 | 2.502 (1.658, 3.774) |
|  | No |  | 24 (20.7) | 92 (79.3) | 116 |  |  |
| Platelets | Thrombocytopenia |  | 14 (60.9) | 9 (39.1) | 23 | 0.005 | 2.006 (1.350, 2.982) |
|  | Normal |  | 54 (30.3) | 124 (69.7) | 178 |  |  |

Neonates with IVH were more likely to die if they were diagnosed with thrombocytopenia (RR=3.306; 95% CI: 1.320, 8.283; p=**0.020**) compared to those with a normal platelet count. Those born in hospital were less likely (RR=0.952; 95% CI: 0.153, 5.910) to die compared to those born at home, however, this relationship was not statistically significant. Vaginal mode of delivery was significantly associated with reduced risk (RR=0.348; 95% CI: 0.122, 0.990; p=**0.031**) of neonatal mortality compared to caesarean sections. Neonates on mechanical ventilation, having a low APGAR score (<7) and with an extremely low birthweight were not significantly associated with mortality (Table 4).

**Table 4:** Factors Associated with neonatal mortality in IVH.

|  |  | Mortality |  | Total | p-value | RR (95% CI:) |
| --- | --- | --- | --- | --- | --- | --- |
|  |  | Died | Survived |  |  |  |
| Gestational Age | Extreme/Very Preterm | 6 (12.2) | 8 (5.7) | 14 | **0.036** | 0.332 (0.128, 0.862) |
|  | Moderate Preterm | 7 (13.0) | 47 (87.0) | 54 |  |  |
| Place of Delivery | Hospital | 12 (19.0) | 51 (81.0) | 63 | 0.958 | 0.952 (0.153, 5.910) |
|  | Home | 1 (20.0) | 4 (80.0) | 5 |  |  |
| Mode of Delivery | Vaginal | 6 (12.0) | 44 (88.0) | 50 | **0.031** | 0.348 (0.122, 0.990) |
|  | Caesarian Section | 7 (38.9) | 11 (61.1) | 18 |  |  |
| APGAR | Low (<=7) | 5 (19.2) | 21 (80.8) | 26 | 0.771 | 1.166 (0.397, 3.424) |
|  | High (>7) | 8 (21.1) | 30 (78.9) | 38 |  |  |
| Mechanical Ventilation | Yes | 9 (20.5) | 35 (79.5) | 44 | 0.709 | 1.227 (0.422, 3.569) |
|  | No | 4 (16.7) | 20 (83.3) | 24 |  |  |
| Platelets | Thrombocytopenia | 6 (42.9) | 8 (57.1) | 14 | **0.020** | 3.306 (1.320, 8.283) |
|  | Normal | 7 (13.0) | 47 (87.0) | 54 |  |  |
| Birthweight | ELBW (<1000g) | 3 (16.7) | 15 (83.3) | 18 | 0.758 | 0.853 (0.258, 2.691) |
|  | VLBW (1000-1500g) | 10 (20.0) | 40 (80.0) | 50 |  |  |

## Discussion

This study reports an IVH prevalence of 33.8% in a tertiary and teaching hospital in Western Kenya where majority of preterm newborns diagnosed with the condition are referred for management. Despite the mounting socioeconomic pressure intraventricular hemorrhage puts in Kenya, this is one of the first studies done to increase the understanding of the condition and its implications on this ever-growing high-risk population.

The prevalence finding is similar to that established in other low-income countries, such as Zambia [14] and Uganda [15] with prevalence being slightly above a third in preterm neonates in both studies. This similarity could be because these studies were carried out in similar setting (LMIC) where resources are limited with need to improve perinatal care to put in measures to prevent occurrence of IVH in the preterm population.

This IVH prevalence finding is lower than that reported in a Polish prospective study [16] among 108 newborns studied, a difference that could be attributed to a lower sample size in the Poznan University of Medical Sciences teaching hospital study. Although both studies were conducted at a teaching hospital, when a lower sample size is adopted, there is a greater likelihood of having a higher proportion of the factor of interest compared to findings in larger hospital or community-based studies conducted over a longer period. In a much larger retrospective study (N=3772) conducted in Canada [17], the prevalence of IVH was estimated at 8.3% which is much lower than the current study. This lower incidence could be attributed to both temporal and methodological differences in the two studies under consideration. The Canadian study [17] reviewed newborns admitted to seventeen neonatal intensive care units (NICUs) between the year 1996 and 1997 while this study was conducted between 2020 and 2021. Disease incidence and prevalence patterns have been demonstrated to vary over time and geographies. Furthermore, reviewing retrospectively findings in multiple facilities with a larger study population could lead to differences in proportions. Furthermore, the authors [17] noted a methodological limitation in their neuroimaging adoption rates across the multiple NICUs studied.

This study adopted the Papile’s grading system [18,19] where Grade I represented subependymal haemorrhage, Grade II - intraventricular haemorrhage without dilation, Grade III -intraventricular haemorrhage with ventricular dilation and Grade IV - intraventricular haemorrhage with parenchymal haemorrhage; the most prevalent grade was Grade I at 51.4% with equal proportions for Grades II, III and IV at 16.2% each. This grade I finding, although lower than that reported in a study conducted in China [4] at 63.2%, shows a similar trend in the highest proportion by IVH grade. However, there were contrasting findings in Grade II and III/IV at 12.3% and 24.1% respectively, a difference that could be attributed to the retrospective design of the study compared to the current study which adopted a prospective study design. Retrospective studies have been fraught with incomplete data and this could explain the merging of grades III and IV by the authors [4]. A second contrasting study in IVH grade proportions was the Polish study where the most prevalent grade reported by the authors [16] was Grade II at 37.3% followed by grade I at 21.6%.

This study identified maternal age, antenatal use of steroids by the expectant mother, mode of delivery (vaginal versus caesarean section), place of delivery (hospital or outborn), gestation age at birth, birthweight, and the neonate’s gender as the probable risk factors for IVH. Use of antenatal steroids significantly reduced the risk of IVH (RR=0.111; 95% CI: 0.028, 0.436; p<0.001) while extreme and very low gestational age below 32 weeks (RR=1.968; 95% CI: 1.255, 3.087; p=0.002) as well as an extremely low birthweight (RR=3.448; 95% CI:2.662, 4.467; p<0.001) significantly increased the risk of IVH two and three-fold respectively. Furthermore, neonates on mechanical ventilation (RR=2.502; 95% CI: 1.658, 3.774; p<0.001) and had thrombocytopenia (RR=2.006; 95% CI: 1.350, 2.982; p=0.005) were significantly more likely to be diagnosed with IVH.

These findings match those of studies conducted in various countries. In Poland [16], having a low gestational age (p=0.0003) and extremely low birthweight (p=0.0007) significantly increased the risk of an IVH diagnosis. Similar to these findings, in Canada [17] gestational age of 27 to 28 weeks increased the risk of IVH two-fold (RR=2.2; 95% CI: 1.5, 3.1) compared to those aged 29 to 32 weeks. In France [20], the authors noted that gestational age (p=0.031) of less than 30 weeks and lower percentile birthweight (p=0.0047) were significantly associated with IVH.

Lastly, this study reports a mortality rate of 19.12% among newborns with IVH. This low mortality rate could be attributed to a high infant survival rate in Kenya following interventions aimed at reducing preterm mortality rate from more than 50% in 2004 [21] to about 33% in 2019 [8]. Kenya is one of the most populous and developed countries in Sub-Saharan Africa and has witnessed an increased investment in healthcare infrastructure that has resulted in a remarkable increase in the survival rates of premature infants [22,23]. However, this improvement in access and quality of care has also caused the significant surge of complications related to preterm birth such as IVH in the country [1]. The study identified gestational age, place of delivery, use of antenatal steroids, mode of delivery, APGAR, birthweight and gender as the probable risk factors for death among newborns diagnosed with IVH. Of these thrombocytopenia (RR=3.306; 95% CI: 1.320, 8.283; p=**0.020**) was significantly associated with mortality. This is in contrast to the findings reported in Poland [16] where gestational age (p=0.0003), low 5-minute APGAR (p=0.00013) and very low birthweight (p=0.0007) were statistically associated with mortality. In the current study, neonates with extremely or very low gestational age (<32 weeks gestation) were significantly more likely to survive (RR=0.332; 95% CI: 0.128, 0.862; p=0.036) compared to those with a moderately low gestation age (32-34 weeks). This could be attributed to the fact that three quarters (75.5%) of these extremely or very low gestational age newborns were on mechanical ventilation compared to 36.8% of those with a moderately low gestation age.

In a study conducted in France [20], a low gestational age at birth (p=0.002) was significantly associated with mortality while use of antenatal steroids was not (p=0.230). In the current study, none of the newborns who mother used antenatal steroids died in the perinatal period. The increased likelihood of survival among neonates born of mothers who had taken antenatal steroids (OR=1.245; 95% CI: 1.105, 1.403) matches the findings reported in a Cochrane review [2] where treatment with antenatal corticosteroids (compared with placebo or no treatment) is associated with a reduction in the most serious pregnancy related adverse outcomes such as prematurity and perinatal death. Corticosteroids use antenatally accelerate fetal lung maturation for women at risk of preterm birth [2].

In Canada [17], vaginal mode of delivery (OR=1.6; 95% CI: 1.2-2.0) and higher (>7) 5-minute APGAR (OR=1.6; 95% CI: 1.2-2.0) scores increased the likelihood of neonatal mortality among those diagnosed with IVH which is in contrast with the current study where vaginal delivery was protective. This difference could be attributed to the difference in the proportionate uptake of vaginal delivery in countries with developing economies and those with developed economies. Higher vaginal deliveries are reporting in these developing economies as most births are supervised and conducted by midwives -who cannot offer caesarean section services – compared to the higher obstetrician to patient ratios in developed countries such as Canada. Finally, this study is limited by the fact that it did not follow-up on the early outcomes of neonates without intraventricular hemorrhage to determine its causality.

## Conclusions and Recommendations

We report an intraventricular haemorrhage incidence among one-third of preterm neonates admitted at a teaching hospital in Western Kenya. Majority of the neonates with intraventricular haemorrhage presented with lower grades of the condition. Their risk of being diagnosed with the condition significantly increased among neonates with very low gestational age and extremely low birthweight; however, it was significantly lower among those neonates whose mothers had taken antenatal steroids. Mortality rates were significantly higher among neonates with both intraventricular haemorrage and thrombocytopenia. We recommend enhanced antenatal steroid use among mothers at risk of having preterm deliveries and newborns with intraventricular hemorrhage. Future studies should follow-up both neonates with and without intraventricular hemorrhage to determine causality.

## Data Availability

All relevant data are within the manuscript and its Supporting Information files.

## Acknowledgements

The authors would like to thank the mothers and the neonates who participated in this study. Secondly, we are grateful to the management of Moi University School of Medicine and Moi Teaching and Referral Hospital for allowing us to use their facilities to conduct the study.

